# Prevalence of IgG antibodies against SARS-CoV-2 among healthcare workers in a tertiary pediatric hospital in Poland

**DOI:** 10.1101/2020.11.28.20239848

**Authors:** Beata Kasztelewicz, Katarzyna Janiszewska, Julia Burzyńska, Emilia Szydłowska, Marek Migdał, Katarzyna Dzierżanowska-Fangrat

**Author notes:** Corresponding author (BK).

## Abstract

Data on prevalence of SARS-CoV-2 antibody in healthcare workers (HCWs) is scare, especially in pediatric settings. The purpose of this study was to evaluate the SARS-CoV-2 IgG-positivity among HCWs of a tertiary pediatric hospital. In addition, follow-up of serological response in the subgroup of seropositive HCWs was performed, to get some insight on persistence of IgG antibodies to SARS-CoV-2. Free, voluntary SARS-CoV-2 IgG testing was made available to HCWs of the Children’s Memorial Health Institute in Warsaw (Poland). Plasma samples were collected between July 1 and August 9, 2020 and tested using the Abbott SARS-CoV-2 IgG assay. Of 2282 eligible participants, 1879 (82.3%) HCWs volunteered to undergo testing. Sixteen HCWs tested positive for SARS-CoV-2 IgG, corresponding to the seroprevalence of 0.85%. Among seropositive HCWs, three had confirmed COVID-19. Of note, 8 (50%) seropositive HCWs reported neither symptoms nor unprotected contact with confirmed SARS-CoV-2 cases in the previous months. A decline in the IgG index was observed at median time of 86.5 days (range:84-128 days) after symptom onset or RT-PCR testing. The nationwide public health response measures together with infection prevention and control practices implemented at the hospital level, at the beginning of the COVID-19 pandemic, might explain a low seroprevalence. Further studies are warranted to elucidate the duration of anti-SARS-CoV-2 antibodies, as well as the correlation between seropositivity and protective immunity against reinfection. Regardless of the persistence of antibodies and their protective properties, such low prevalence indicates that this population is vulnerable to a second wave of the COVID-19 pandemic.

## Introduction

Severe acute respiratory syndrome coronavirus 2 (SARS-CoV-2), causing coronavirus disease 2019 (COVID-19) which emerged in December 2019, has evolved to a global pandemic [1]. In Poland first, imported COVID-19 case was reported on March, 3 2020 and 3 weeks later a nationwide lockdown was commenced [2]. Until August, 31, there were 66 870 confirmed cases, with 2 033 COVID-19 related deaths [3].

In the Masovian district (one of the three most affected regions in Poland) the first cases were recorded on March 13 and by the end of August 2020, there were 9370 cases and 411 deaths [4].

Although real-time RT-PCR is considered the gold standard for the diagnosis of the acute SARS-CoV-2 infection, this test is limited by transient nature of RNA. In addition, the sensitivity of RT-PCR methods is estimated to be no higher than 70% [5], which may lead to underdiagnosing of SARS-CoV-2 infections, especially in subclinical or asymptomatic cases. By identifying individuals who have developed antibodies to the virus (including those that may be asymptomatic or have recovered), serology can give greater details into the prevalence of SARS-CoV-2. Although, concerns have arisen on persistence of IgG antibodies to SARS-CoV-2 after recovery [6,7].

Two entitles of infected individuals pose the highest risk for SARS-CoV-2 transmission in the hospital setting. First, infected patients until diagnosis. Second, SARS-CoV-2-positive health care workers (HCWs). As children and adolescent comprise less than 5% of all positive cases in Europe [8], majority of SARS-CoV-2 infections among HCWs in pediatric hospitals might be associated with transmission in community or from infected co-workers.

Data on SARS-CoV-2 prevalence among HCWs in pediatric hospital settings is scare [9,10].

Knowing the prevalence of SARS-CoV-2 infection among HCWs is vital to inform pandemic response. The aim of this study was to evaluate the SARS-CoV-2 IgG positivity among HCWs of a tertiary pediatric hospital in Warsaw (Masovian district), Poland. In addition, we have performed follow-up of serological response to SARS-CoV-2 in the subgroup of seropositive HCWs, to get some insight on persistence of specific antibodies.

## Materials and methods

Free, voluntary SARS-CoV-2 IgG testing was made available to the hospital staff of the Children’s Memorial Health Institute (CMHI) in Warsaw, Poland (including physicians, nurses, other workers with direct patient contact, i.e. physical therapists as well as workers without direct patient contact, i.e. laboratory workers, pharmacists, administrative staff, maintenance, etc.). All participants were asymptomatic at the time of serology testing. In particular, those who were previously symptomatic, had no symptoms for at least 14 days. Plasma (EDTA) samples were collected between July 1 and August 9, 2020 (corresponding to 97 – 135 days after the nation-wide lockdown was commenced).

Plasma samples were run on the Abbott Alinity i instrument using the Abbott SARS-CoV-2 IgG assay (Abbott Laboratories, Lake Bluff, IL, USA) following manufacturer’s instruction. The assay is a chemiluminescent microparticle immunoassay (CMIA) for qualitative detection of IgG antibodies to the nucleocapsid (N) protein of SARS-CoV-2. The manufacturer’s index value (a signal/cut-off; S/CO ratio) of ≥1.40 was interpreted as positive. The assay has been shown to have 99.9% specificity and 100% sensitivity for samples taken greater than 17 days post symptom onset [11].

Demographic data (age, gender), results of SARS-CoV-2 RNA testing (if performed in CMHI) were collected for all participants, from the laboratory records. Data on profession and the necessity of quarantine or isolation (date and duration of quarantine or isolation) were collected from human resource’s database. In addition, seropositive or previously SARS-CoV-2 infected individuals were approached telephonically and data regarding contacts with a confirmed or suspected COVID-19 case, the positive test result in the past (if performed outside CMHI), the necessity of inpatient treatment, experienced symptoms over the previous months, were collected by the head of the infection control department. All data were analyzed anonymously.

### Study setting

The CMHI in Warsaw (Masovian district), is the largest tertiary pediatric hospital and research institute in Poland. With over 590 beds and 2282 employees it performs over 249 000 services (including inpatients and outpatients) per year.

At the time of serology testing, we had no case of SARS-CoV-2 infection among patients. Until July1, 2020, the first day of the serology testing, we had had 5 confirmed SARS-CoV-2 infections among HCWs (all contracted outside the hospital setting, one confirmed outside CMHI) and there were no additional cases until July 6, 2020. Since that time until August 9, 2020 (i.e. the end of the serology testing), additional 4 linked cases among laboratory staff, were confirmed by RT-PCR (**Fig 1**).

**Figure 1.**
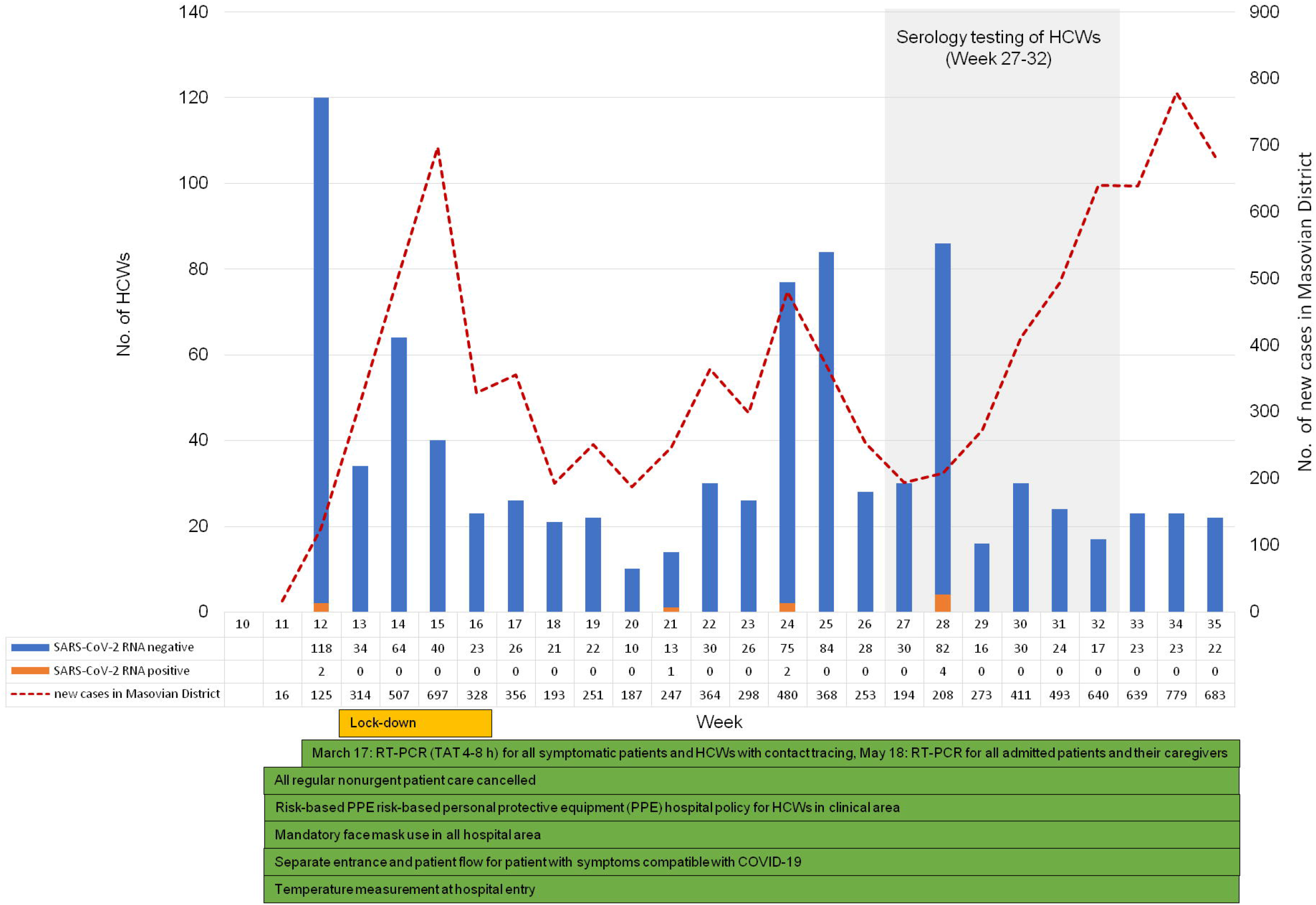
The epidemic curve is shown as the number of HCWs tested each week for SARS-CoV-2 RNA by RT-PCR in CMHI together with the number of new SARS-CoV-2 cases recorded in the Masovian district. The first case of SARS-CoV-2 RNA among HCWs of CMHI was detected on March, 17 2020. Voluntary serological testing for HCWs of CMHI was conducted from July 1 to August 9, 2020 (corresponding to week 27 and 32). Details of infection prevention and control measures implemented at CMHI together with the nationwide lockdown, are given below the curve.

### Statistical analysis

Statistical analysis was carried out using the Statistica data analysis software system (TIBCO Software Inc.), version 13. Continuous variables were presented as median and interquartile range (IQR). Categorical variables were summarized using percentages and counts. Seroprevalence of SARS-CoV-2 IgG was calculated as proportion with 95% confidence intervals (CI). The association between variables was tested with Chi-square or Fisher’s exact test (for categorical variables) and Mann Whitney U test (for continuous variables). Univariable and multivariable logistic regression analysis were run to evaluate factors associated with seroprevalence of SARS-CoV-2 IgG. For the variables to be included in multiple logistic model, a stepwise selection was used, starting with the full model, and using p-value of 0.1 for removal and 0.05 for addition of variables.

### Ethical consideration

This study reporting the results of free-voluntary serology testing, was not offered as a research protocol but as a service to healthcare workers. The study has been reviewed and approved by the Institutional Review Board of the Children’s Memorial Health Institute in Warsaw (Ref. no. 10/P-IN/20), and granted a waiver of consent since the data were analyzed anonymously.

## Results

### Baseline characteristics

Of 2282 eligible participants, 1879 HCWs volunteered to undergo testing, yielding a participation rate of 82.3%. Median (IQR) age was 48 (38-56) years, and 85.8% were female. Approximately one third (639/1879, 34%) were nurses, and 19.7% were physicians.

Majority (70.9%) of HCWs worked in the clinical area. Twenty-two per cent (417/1879) had been tested for SARS-CoV-2 RNA by RT-PCR as a part of implemented infection control measurements (note that administration of RT-PCR test does not imply that a HCW was suspected of having COVID-19, since tests were also performed as a part of contact tracing, in previously quarantined or isolated HCWs before return to work or in newly employed staff). The median time between RT-PCR and serology testing was 77 days (IQR: 39-122 days). SARS-CoV-2 RNA was detected in 4 of 417 HCWs, including 3 HCWs with positive serology (details given below). The remaining one, with a positive SARS-CoV-2 RNA test, was seronegative. This was the HCW whose plasma was collected 129 days post RT-PCR testing.

Fifty-six HCWs had been quarantined or isolated. The median time since the start of quarantine or isolation to serology testing was 97.5 days (IQR: 42.5-126 days) (**Table 1**).

**Table 1.** Baseline characteristics of 1879 HCWs.

| Characteristics | Total (%) |
| --- | --- |
| Sex: |  |
| male | 266 (14.2) |
| female | 1613 (85.8) |
| Age, median (IQR) years | 48 (38-56) |
| Professional category:<sup>a</sup> |  |
| nurse | 639 (34.0) |
| physician | 371 (19.7) |
| other with a direct patient contact | 226 (12.0) |
| other without a direct patient contact | 643 (34.2) |
| Healthcare department:<sup>b</sup> |  |
| clinical | 1332 (70.9) |
| non-clinical | 547 (29.1) |
| Previously tested for SARS-CoV-2 RNA |  |
| yes<sup>c</sup> | 417 (22.2) |
| no | 1462 (77.8) |
| Quarantined or isolated: |  |
| yes<sup>d</sup> | 56 (3.0) |
| no | 1823 (97.0) |

<sup>a</sup>*other with a direct patient contact*: patient care technician (67); physical therapist (43), radiation therapist (37); psychologist (21), medical assistant (11), audiologist (10), pedagogue (9), speech pathologist (8), clinic engineer (6), medical technician (6), dentist assistant (4), anthropologist (4); *other without a patient contact*: office worker (228), secretary (136), laboratory worker (110), kitchen worker (34), pharmacist (32), dietician (27), medical sterilization technician (19); driver (17), labourer (15), store man (8), IT worker (7), public health worker (3), manager (2); security officer (2), chaplain (2), parking attendant (1).
<sup>b</sup>*clinical departments* include: medical (765), surgical (222), auxiliary medical (123), ambulatory (116), intensive care (106); *non-clinical departments* include: administration (284), laboratory (113), maintenance (116), pharmacy (34).
<sup>c</sup> median period between SARS-CoV-2 RNA and serology testing was 77 days (range 1 – 136 days; IQR 39-122 days); 4 out of 417 (0.96) HCWs had positive test results.
<sup>d</sup> median period between the start of quarantine/isolation and serology testing was 97.5 days (range 10 -133 days; IQR 42.5-126 days).

### Seroprevalence among HCWs

Sixteen healthcare workers tested positive for SARS-CoV-2 IgG, corresponding to the seroprevalence of 0.85%. Median index was 2.69 S/CO (range: 1.41-7.59 S/CO).

Of the 16 seropositive HCWs, only 3 (18.75%) tested positive for SAR-CoV-2 RNA (within the prior 53-106 days). Five seropositive HCWs were RT-PCR negative within the prior 6-127 days (only one of them had been tested less than 14 days prior to serology). The remaining eight seropositive HCWs had not been tested by RT-PCR (all of them were negative, when tested 1-11 days after IgG testing).

Among seropositive HCWs, six (37.5%) presented symptoms compatible with COVID-19, one had a household contact with a suspected COVID-19 case, whereas 8 (50%) reported neither symptoms compatible with COVID-19 in the prior months nor unprotected close contacts with confirmed or suspected COVID-19 cases (**Table 2**).

**Table 2.** Characteristics of the seropositive HCWs.

| No. | Age (years) | Gender | SARS-CoV-2 IgG (S/CO) | SARS-CoV-2 RNA by RT-PCR | Time since RT-PCR testing and serology testing | Symptoms compatible with COVID-19 | Possible route of SARS-CoV-2 transmission |
| --- | --- | --- | --- | --- | --- | --- | --- |
| 1 | 50 | f | 3.07 | negative | 2 days* | no | unknown |
| 2 | 45 | f | 1.85 | negative | 2 days* | no | unknown |
| 3 | 60 | f | 1.52 | negative | 11 days* | no | unknown |
| 4 | 60 | f | 1.41 | negative | 6 days | no | unknown |
| 5 | 60 | f | 5.00 | positive | 43 days | yes | household contact with a confirmed case |
| 6 | 50 | f | 1.66 | negative | 3 days* | no | unknown |
| 7 | 50 | f | 6.92 | negative | 1 day* | no | household contact with a suspected case |
| 8 | 45 | f | 2.04 | (multiple) negative | 14, 34, 42, 111 days | yes | unknown |
| 9 | 40 | f | 2.42 | (multiple) negative | 62, 127 days | no | unknown |
| 10 | 55 | m | 2.86 | positive | 106 days | yes | unknown |
| 11 | 55 | f | 4.32 | negative | 82 days | no | unknown |
| 12 | 40 | f | 2.51 | negative | within 24 h* | no | unknown |
| 13 | 50 | f | 3.76 | positive | 53 days | yes | unknown |
| 14 | 60 | f | 7.59 | negative | 14 days | yes | unknown |
| 15 | 65 | m | 2.21 | negative | 2 days* | no | unknown |
| 16 | 50 | f | 7.32 | negative | within 24 h | yes | unknown |

157 \* indicates HCWs who were tested by RT-PCR after serology testing results were obtained.

### Factors associated with SARS-CoV-2 IgG positivity

The odds of being seropositive were higher in HCWs who had been previously tested by RT-PCR regardless of the test results (adjusted OR=3.82, 95% CI: 1.42-10.29; p=0.008) (**Table 3**). The age, sex, professional category or working in clinical area, did not show any statistically significant association with the positivity for SARS-CoV-2 IgG (**Table 3**).

**Table 3.** Analysis of factors associated with SARS-CoV-2 IgG positivity.

| Characteristics | SARS-CoV-2 IgG |  | P-value<sup>a</sup> | Univariate |  | Multivariate |  |
| --- | --- | --- | --- | --- | --- | --- | --- |
|  | negative | positive |  | OR (95% CI) | P-value | OR (95% CI) | P-value<sup>c</sup> |
| Sex: |  |  | 0.849 |  | 0.849 |  |  |
| male | 264 (14.2) | 2 (12.5) |  | 1 |  |  |  |
| female | 1599 (85.8) | 14 (87.5) |  | 1.16 (0.26 – 5.11) |  |  |  |
| Age, median (IQR) years<sup>b</sup> | 48 (38-56) | 50.5 (46-57.5) | 0.141 | 1.03 (0.99 – 1.08) | 0.131 | 1.04 (0.99 – 1.09) | 0.093 |
| Professional category: |  |  | 0.114 |  |  |  |  |
| other without a direct patient contact | 639 (34) | 4 (25) |  | 1 |  |  |  |
| nurse | 635 (34) | 4 (25) |  | 1.01 (0.25 – 4.04) | 0.992 |  |  |
| physician | 364 (20) | 7 (44) |  | 3.07 (0.89 – 10.56) | 0.075 |  |  |
| other with a direct patient contact | 225 (12) | 1 (6) |  | 0.71 (0.08 – 6.39) | 0.760 |  |  |
| Healthcare department: |  |  | 0.716 |  | 0.717 |  |  |
| non-clinical | 543 (29) | 4 (25) |  | 1 |  |  |  |
| clinical | 1320 (71) | 12 (75) |  | 1.23 (0.40 – 3.84) |  |  |  |
| Previously tested for SARS-CoV-2 RNA |  |  | 0.007 |  | 0.012 |  | 0.008 |
| no | 1454 (78.05) | 8 (50) |  | 1 |  |  |  |
| yes | 409 (21.95) | 8 (50) |  | 3.56 (1.33 – 9.53) |  | 3.82 (1.42-10.29) |  |

<sup>a</sup>Chi-squared test<sup>b</sup>Mann Whitney U test<sup>c</sup>Wald test

### Follow up serological data

Three (i.e. no. 5, 11 and 14) out of 16 seropositive HCWs provided second sample for serology testing. Time span between initial testing and the second sample was 36 (no.11), 57 (no. 14) and 85 (no. 5) days, which corresponds to 88, 118 and 128 days, post symptom onset (no. 5 and 14) or RT-PCR testing (no. 11). Decrease in SARS-CoV-2 IgG index value was observed in all 3 cases.

In addition, we monitored three initially seronegative HCWs, diagnosed with COVID-19 within a week following their first serology testing (i.e. in the week 27). All three were epidemiologically linked and thus considered clustered cases with intra-hospital transmission. These HCWs were immediately put on home isolation as soon as they were identified by RT-PCR testing (all experienced mild symptoms). After their return to work, SARS-CoV-2 IgG was tested twice in convalescent plasma samples collected 34-37 and 84-85 days after the RT-PCR testing. Decrease in the index value were observed in all 3 cases between the second and the third month after symptom onset.

Overall, all six HCWs remained seropositive while tested on the last sample collected at the median time of 86.5 days (range: 84-128 days) after symptom onset or RT-PCR testing (**Fig 2**).

**Figure 2.**
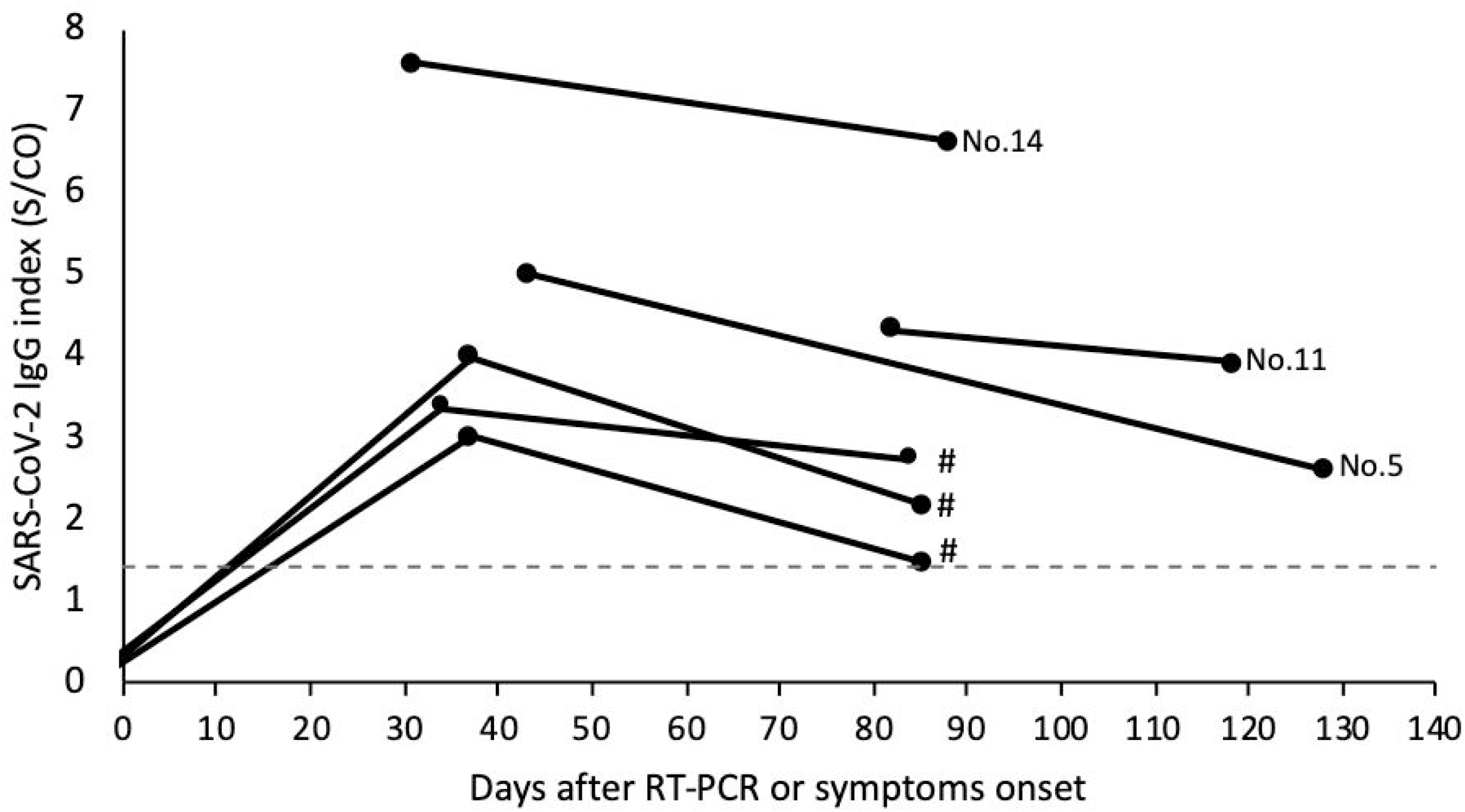
SARS-CoV-2 IgG follow-up data in 6 HCWs including three seropositive at initial testing (no. 5, 11 and 14) and three HCWs clustered cases (indicated by #), who had confirmed COVID-19 within a week following initial serology testing (in the week 27). The dashed line indicates positivity threshold (1.4 S/CO).

## Discussion

In Poland, a nationwide lockdown, including closure of school, public institutions, and mandatory face-mask was implemented during the very early stages of the COVID-19 pandemic [2]. As a result, the daily numbers of new SARS-CoV-2 infections have been relatively stable up to the study period with cumulative incidence of 90.6 per 100 000 as on July 1 2020. Preceding nationwide public health response measures, a set of infection prevention and control measures had been implemented in CMHI, to contain the spread of SARS-CoV-2 infection within the hospital. These included (but were not limited to): measurement of body temperature at the hospital entry, patient/caretaker screening for SARS-CoV-2 RNA by RT-PCR on nasopharyngeal swabs on admission and RT-PCR testing of all symptomatic staff coupled with contact tracing.

Seroprevalence studies in HCWs allow for assessing the level of exposure, and indirectly, the effectiveness of implemented protective measures. They are also crucial to inform health care resource planning to provide safe environment to protect both patients and HCWs from SARS-CoV-2 infection. A few seroprevalence studies of HCWs in pediatric hospitals have been published so far. Here, we present the results of serology testing in HCWs of the large pediatric hospital in Poland, including over 82% of employees. In our study SARS-CoV-2 IgG positivity rate was 0.85%, and it was lower than in previous reports from pediatric hospitals. The seroprevalence of SARS-CoV-2 among 175 HCWs in a large referral pediatric hospital in Barcelona was 4% [10]. Another study form Italy, performed at the same time (i.e. mid-April 2020), showed seroprevalence of 5.13% [12]. Timing of serology testing (April 2020 *vs* July 2020) and significantly higher burden of COVID-19 (as Spain and Italy were two most affected countries in Europe, [13]) may account for this difference. In addition, and in contrast to Spanish and Italian reports, we did not have any confirmed COVID-19 case among patients in our hospital until the end of the study period.

Interestingly, we did not observe any difference in seroprevalence between clinical and non-clinical working locations or across professional groups. This may be due to the fact that at that time we did not managed any children with confirmed COVID-19. Although asymptomatic SARS-CoV-2 carriage among hospitalized children cannot be completely ruled out (since RT-PCR screening on admission is not 100% sensitive to preclude infection), the risk of children to staff transmission seems to be low. The recent study from pediatric hospital in Chicago, revealed a low (1-2 %) prevalence of SARS-CoV-2 among children without COVID-19 symptoms as well as no secondary transmission among HCWs exposed to these patients [14]. Another study, comparing dynamics and determinants of SARS-CoV-2 transmissions among HCWs of adult and pediatric settings in Paris, revealed significantly lower attack rate in the pediatric setting (2.3% *vs* 3.2%, respectively) [15].

In our study, prior RT-PCR testing regardless of the result, was associated with increased adjusted risk of SARS-CoV-2 IgG positivity. This may be a reflection of the fact that, testing for SARS-CoV-2 RNA was performed as a part of contact tracing and in previously quarantined or isolated HCWs before return to work. Considering that as many as 50% of seropositive HCWs in our study were asymptomatic or had no confirmed contact with suspected or proven COVID-19 case, and that over 80% (13/16) had not been tested or tested negative for SARS-CoV-2 RNA, it could indicate that some SARS-CoV-2 infections among HCWs were unrecognized or undetected. Although, false-positive SARS-CoV-2 IgG results are possible (e.g. due to cross-reactivity to commonly circulating human coronaviruses) they are unlikely, even in the limited circulation of the virus [11]. Thus, it seems reasonable to test both symptomatic and asymptomatic HCWs for SARS-CoV-2 RNA on a regular basis, if resources are available.

We evaluated antibody persistence in a subgroup of HCWs with multiple plasma samples available for serology testing, to get some insight into antibody persistence. SARS-CoV-2 IgG antibodies were detected up to 128 days post symptoms onset or RT-PCR testing. However, the index value declined consistently in all subjects between first and last plasma sample tested. Our findings are in line with some previously published data on kinetics of SARS-CoV-2 IgG [16,17]. Recent study by Strömer et al. evaluated SARS-CoV-2 IgG levels in follow-up samples from 16 individuals (median time of the last sample submission was 153 days after the RT-PCR) and revealed that several SARS-CoV-2 infected patients lost their N-specific IgG within a few months or could lose them soon [16]. Of note, in contrast to N-specific IgG, the response to the spike (S) protein was found to be more stable and was associated with the presence of virus-neutralizing antibodies (although at relatively low titres). Another study by Patel et al., evaluated change in antibodies to SARS-CoV-2 over 60 days among 19 HCWs (including symptomatic ones), using S-based assay [17]. They observed a decrease in anti-SARS-CoV-2 antibodies in all HCWs, with 58% of seropositive individuals becoming seronegative. Taking together, these findings suggest that seroprevalence studies may underestimate rates of prior infections as antibodies may only be transiently detectable after infection. Nevertheless, the limited number of HCWs with follow-up sample available precludes any meaningful conclusion. In contrast to ours and two studies mentioned above, a recent population-based study, implementing two highly sensitive and specific assays to monitor antibody levels and their durability, indicated that antibody remained stable over 4 months. These discordant results may be attributed to sampling biases [18]. Thus, larger longitudinal serological studies are warranted, including these with virus neutralization assays, to explore the dynamic and the persistence of the SARS-CoV-2 antibodies as well as their correlation with immunoprotection from reinfection.

Our study has some limitations which should be acknowledged. First, testing spanned an over a 5-week period (week 27 – 32), potentially leading to changes in incidence over time and possible variation in professional groups attending testing. Second, we cannot rule out that some of HCWs were infected and either mounted no detectable antibody response or it had weaned by the time of serology testing, as we had found one of confirmed COVID-19 case seronegative 129 days after the positive RT-PCR test. Thus, the seroprevalence in our study could be underestimated. Moreover, data on symptoms, exposure histories or personal protective equipment use were collected only for the subset of seropositive HCWs by telephone interview (subjected to recall bias), therefore more detailed information on risk factors could not be assessed. Nevertheless, so far this is the largest study assessing the prevalence of SARS-CoV-2 IgG antibodies among HCWs in the pediatric hospital setting, with a high response rate and the use of high-quality serological assay. Our study provides data on the seroprevalence of SARS-CoV-2 infection among HCWs in the pediatric setting in the initial peak of the pandemic, which inform control and prevention strategies for future waves of COVID-19 pandemic.

## Conclusions

SARS-CoV-2 seroprevalence in healthcare workers of a tertiary pediatric hospital in Poland is low (0.85%). The nationwide public health response measures together with infection prevention and control practices implemented at the hospital level, at a start of COVID-19 pandemic, might explain a low seroprevalence. Further studies are warranted to elucidate the duration of anti-SARS-CoV-2 antibodies, as well as the correlation between seropositivity and protective immunity against reinfection. Regardless of the persistence of antibodies and they protective properties, such low prevalence indicates that this population is vulnerable to a second wave of COVID-19 pandemic.

## Data Availability

All relevant data will be held in a public repository at https://data.mendeley.com

http://dx.doi.org/10.17632/5jmxvscmgv.1

## Notes

### Competing Interest Statement

The authors have declared no competing interest.

### Author Declarations

The study has been reviewed and approved by the Institutional Review Board of the Children's Memorial Health Institute in Warsaw (Ref. no. 10/P-IN/20), and granted a waiver of consent since the data were analyzed anonymously.

